# Equivalence of plasma and serum for clinical measurement of p-tau217: comparative analyses of four blood-based assays

**DOI:** 10.1101/2024.12.26.24319657

**Authors:** Yijun Chen, Ally L. Albert, Anuradha Sehrawat, Marissa Farinas, Oscar L. Lopez, Xuemei Zeng, Ann D. Cohen, Thomas K. Karikari

## Abstract

**Background:** Phosphorylated tau (p-tau) 217 is a promising blood biomarker for Alzheimer’s disease (AD). However, most p-tau217 assays have been validated solely in ethylenediaminetetraacetic acid (EDTA) plasma, leaving the clinical applicability of serum p-tau217 largely unexplored despite serum being a preferred matrix in many clinical laboratories. To address this gap, we compared p-tau217 concentrations and diagnostic performances in matched plasma and serum samples using four research-use-only assays, including three from commercial sources i.e., Lumipulse, ALZpath, NULISA, and one from University of Pittsburgh.

**Methods:** Paired plasma and serum samples were processed from the same venipuncture collection and assessed with the four p-tau217 assays following manufacturer-recommended procedures in two research cohorts (N=84).

**Results:** Plasma and serum p-tau217 levels varied across assays; the ALZpath, Pittsburgh, and NULISA methods showed significantly lower p-tau217 levels in serum compared with plasma (p<0.0001), while Lumipulse showed higher or non-significant differences in serum. Yet, strong correlations (rho >0.8) were observed between plasma and serum p-tau217 pairs. Both plasma and serum p-tau217 demonstrated strong classification accuracies to differentiate clinical AD from normal controls, with high AUC (up to 0.963) for all methods. The exception was the Pittsburgh assay, where plasma p-tau217 had superior AUC than serum p-tau217 (plasma: 0.912, serum: 0.844). The rest of the assays had equivalent accuracies in both matrices.

**Conclusions:** Serum p-tau217 performs equivalently as plasma p-tau217 for most assessed assays. Serum can therefore be used in place of plasma for p-tau217 assessment for research and clinical purposes.

## Introduction

Biomarkers enable accurate diagnosis, treatment, and research of Alzheimer’s disease (AD). Currently, positron emission tomography (PET) neuroimaging and cerebrospinal fluid (CSF) biomarkers are clinically approved methods for AD diagnosis. [1, 2] However, their invasiveness, high costs, and limited availability hamper applications in routine clinical practice and population screening. [3] In contrast, blood-based biomarker assays offer cost-effective and minimally invasive alternatives, enhancing the accessibility and widespread availability of AD diagnostics. [4, 5]

Phosphorylated tau217 (p-tau217) is one of the most promising AD blood biomarkers available today, recommended for use in the latest diagnostic and research guidelines for the field, including the 2024 Alzheimer’s Association revised criteria [6] and the 2024 International Working Group update. [7] Blood p-tau217 can identify individuals with biological evidence of AD across the disease continuum, and robustly distinguish AD from other neurodegenerative disorders. [8–13] Additionally, plasma p-tau217 levels increase with brain amyloid-beta (Aβ) plaque and tau tangle deposition, allowing for early detection and monitoring of the disease. [14, 15]

Multiple academic and industrial groups have developed plasma-based p-tau217 assays classified under two main techniques: immunoassays and mass spectrometry, with most assays being the former (for a review, see [16]). These p-tau217 assays have been mainly tested in ethylenediaminetetraacetic acid (EDTA) plasma (hereafter called plasma). However, clinical laboratories in many hospital systems preferentially collect serum over plasma. [17] Despite this preference, the utility and clinical applicability of serum p-tau217 remain largely unexplored. Assessing p-tau217 in serum is essential to guide its adoption in such clinical settings.

We compared the levels and predictive performances of matched plasma vs. serum p-tau217 to distinguish between Aβ pathology and cognitive status groups using samples from two distinct cohorts. We examined four research-use-only p-tau217 assays: three from commercial sources (Lumipulse from Fujirebio, ALZpath from Quanterix and the Nucleic acid linked immuno-sandwich assay [NULISA] from Alamar Biosciences) and the fourth from an academic laboratory (University of Pittsburgh).

## Materials and Methods

### 2.1 Participant recruitment

This study included participants from two distinct cohorts. The first cohort was from the University of Pittsburgh Alzheimer’s Disease Research Center (Pitt-ADRC). The participants underwent annual clinical evaluations to monitor brain health and detect signs or progression of cognitive decline. [18, 19] Neuropsychological testing involved a range of cognitive tests, such as the Montreal Cognitive Assessment (MoCA), Mini-Mental State Examination (MMSE), and the Clinical Dementia Rating (CDR). The present investigation was a prospective, blinded sub-study of the Pitt-ADRC cohort focusing on those attending their annual clinical appointments and thus did not have neuroimaging data available.

The second cohort, the Human Connectome Project (HCP) study, is a community-based cohort where participants are subjected to a detailed battery of cognitive tests such as the MoCA, tests of verbal fluency, visual naming, Trail Making Test, verbal free recall, and the Rey-Osterrieth Complex Figure Test to evaluate their cognitive function. Neuroimaging assessments included structural and functional magnetic resonance imaging (MRI), diffusion tensor imaging, and PET imaging with Pittsburgh Compound B (PiB) for brain Aβ deposits. [20]

The ADRC and HCP studies were approved by the University of Pittsburgh Institutional Review Board (MOD19110245-023 and STUDY19100015C, respectively). All participants provided written informed consent, in compliance with the ethical standards and protocols sanctioned by the university’s Human Research Protection Office.

### 2.2 Blood collection and processing procedures

Blood samples were collected via venipuncture by experienced nurses from 9:00 am - 2:00 pm, taking note of the time of last meal. [21]

A 10 ml Lavender top EDTA plasma tube (BD #366643) and a 10 ml red top serum tube (BD #368045) were used to collect whole blood from each participant. The tubes were promptly inverted 8-10 times (EDTA tube) and 5 times (serum tube) incubated at room temperature for 30 minutes and centrifuged at 2000 xg for 10 minutes at 4°C. The resulting plasma and serum samples were aliquoted into cryovials and frozen at −80°C until use.

### 2.3 P-tau217 measurements

The plasma and serum samples were thawed, vortexed, and centrifuged at 4,000 xg for 10 minutes to remove particulates. The clarified supernatants were analyzed using various assays. Identical reagent batches were used for plasma and serum samples measured in duplicates except for NULISA (details below). All measurements were conducted at the Biofluid Biomarker Laboratory, Department of Psychiatry, School of Medicine, University of Pittsburgh, PA, USA. Researchers were blinded to all participant information until the completion of data acquisition.

#### ALZpath p-tau217

The ALZpath-p-tau217 kit (Quanterix, #104570) was used on the Single molecule array (Simoa) HD-X platform, following the manufacturer’s protocols. Three levels of Quality control (QC) samples were analyzed in duplicate at the beginning and the end of each run. The within- and between-run variations for the three QCs ranged between 2.0%-6.5% and 5.8%-9.7%, respectively, for the ADRC cohort and 1.5%-11.5% and 7.6%-17.0% for the HCP cohort. One serum sample failed in the HCP cohort and was excluded from further analysis.

#### Pittsburgh p-tau217

The Pittsburgh assay, developed at the University of Pittsburgh on the Simoa HD-X platform [22, 23], employs a p-tau217 monoclonal antibody coupled to paramagnetic beads (#103207, Quanterix), for capturing the antigen, and a biotin-conjugated mouse monoclonal antibody against the N-terminal region of tau (Tau12; BioLegend, #SIG-39416) for detection. *In vitro* phosphorylated recombinant full-length tau-441 (T08-50FN-50, SignalChem) was used as the calibrator. Samples and calibrators were diluted threefold to position the assay signals around the mid-point of the standard curve. For each run, two levels of QC samples were analyzed in duplicates at the start and end of each run. The within- and between-run variations were 7.1-7.8% and 11.6-17.5% for ADRC and HCP cohorts, respectively. One serum sample failed in the HCP cohort and was excluded from further analysis.

#### Lumipulse p-tau217

The fully automated Lumipulse G1200 was used following the manufacturer’s guidelines. We utilized commercially sourced p-tau217 kits (#81471) from Fujirebio Europe (Ghent, Belgium). Two levels of QCs were measured at both the beginning and end of each batch, and both fell within the manufacture-supplied acceptable concentration ranges.

#### NULISA assay

The NULISAseq CNS disease panel 120 (v2) on an Alamar ARGO^TM^ system was used. [11][24] Briefly, samples were incubated with paired capture and detection antibodies, with mCherry protein as an internal control. Capture antibodies were conjugated to DNA with a poly-A tail and target-specific barcode, while detection antibodies were conjugated to DNA containing a biotin group and complementary barcode. Immunocomplexes were captured on magnetic beads, washed, and released, then recaptured using streptavidin-coated beads. Target- and sample-specific barcoded DNA reporters were synthesized via ligation and quantified using next-generation sequencing. Protein levels were normalized to generate NULISA Protein Quantification (NPQ) units by dividing target counts (plus one) by mCherry counts (plus one). The initial NPQ values were log2 transformed. To make the data more intuitive, we then applied an inverse log2 transformation (2^NPQ) for the analysis. All samples were analyzed in a single run. QC samples were processed following the manufacturer’s guidelines, achieving an intra-plate variation of 18.23%. We focused on p-tau217 data in this study, leaving out the other analytes.

### 2.4 Statistical analysis

For participant demographic characteristics, continuous variables were summarized using median and interquartile range (IQR), while categorical variables were reported as numbers and percentages. Differences between clinical and Aβ PET groups for continuous variables were examined using the Wilcoxon Rank Sum test. Categorical variables were analyzed using Fisher’s exact tests. The Wilcoxon Rank Sum test was also used to assess the discriminating performance of biomarkers on cognitive status for the Pitt-ADRC cohort. Due to the small sample size of the Aβ PET-positive cases in the HCP cohort (only three cases), no comparative statistical test was performed. Spearman correlation analysis was conducted to evaluate the strength of associations among different assay platforms and between plasma and serum. The Delong test was used to compare the area under the curve (AUC) difference between the plasma and the corresponding serum assays. For all tests, a p-value less than 0.05 was considered statistically significant. Analyses were performed using the R statistical software (version 4.2.1, R Foundation for Statistical Computing, Vienna, Austria). Plots were generated either with R or GraphPad Prism (version 10.1.1).

## Results

### Participant characteristics

In the Pitt-ADRC cohort (n=50), the median age was 76 (70.3- 82.0) years, with 21 (42.0%) females. Sixteen (32.0%) were diagnosed with probable AD and thirty-four (68.0%) as normal controls. In terms of cognitive performance, the median MMSE and MoCA scores were 27.0 (23.0- 29.0) and 23.0 (16.5- 27.0), respectively. Sixteen participants (32.0%) had a CDR score of “disease absent” (CDR=0), 24 (48.0%) were scored as “questionable” (CDR=0.5), and 5 (10.0%) each as “disease present but mild” (CDR=1), and “moderate” (CDR=2). There were significant differences in MoCA, MMSE and CDR scores between the probable AD and the normal control groups. (Table 1)

**Table 1.**
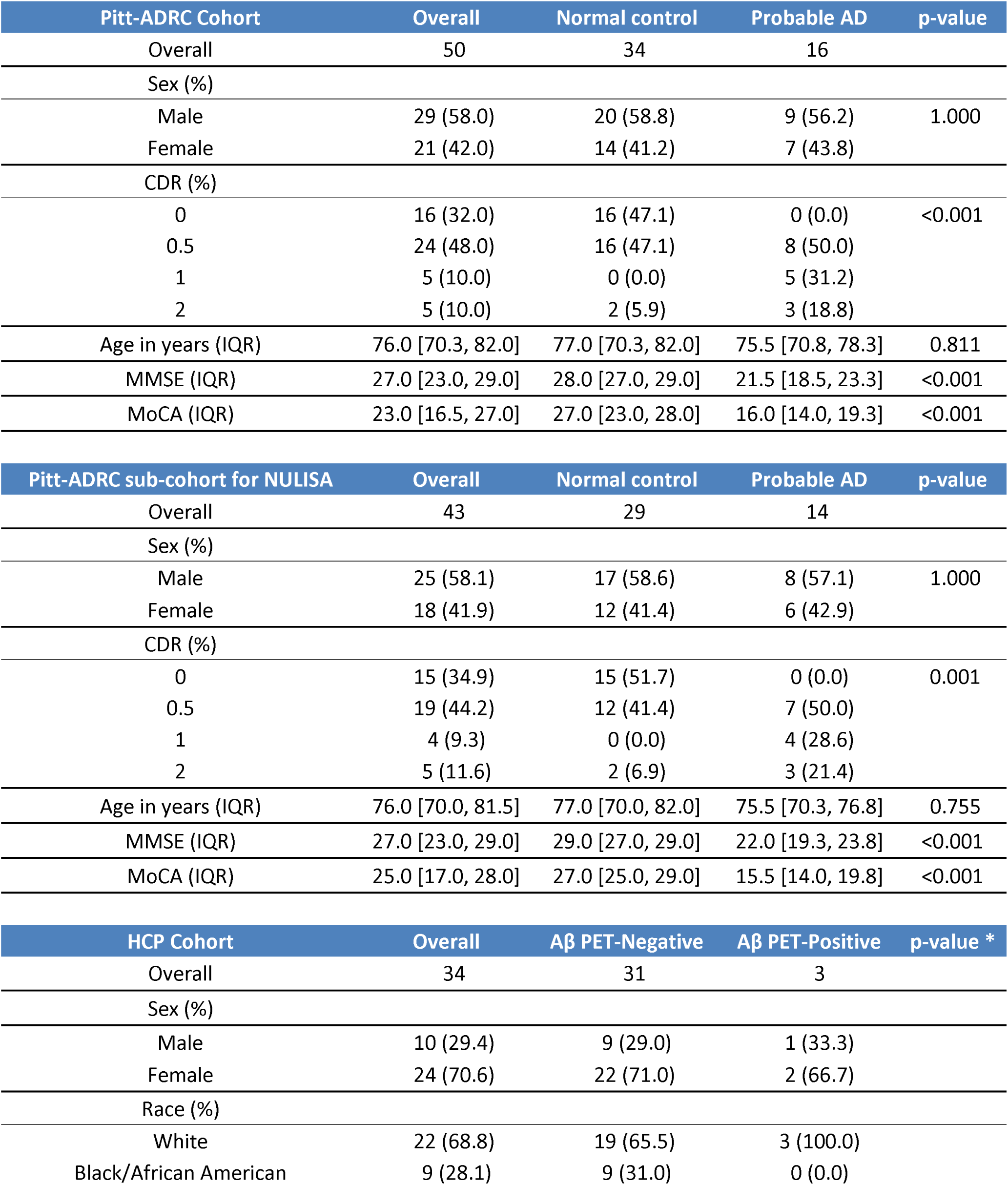
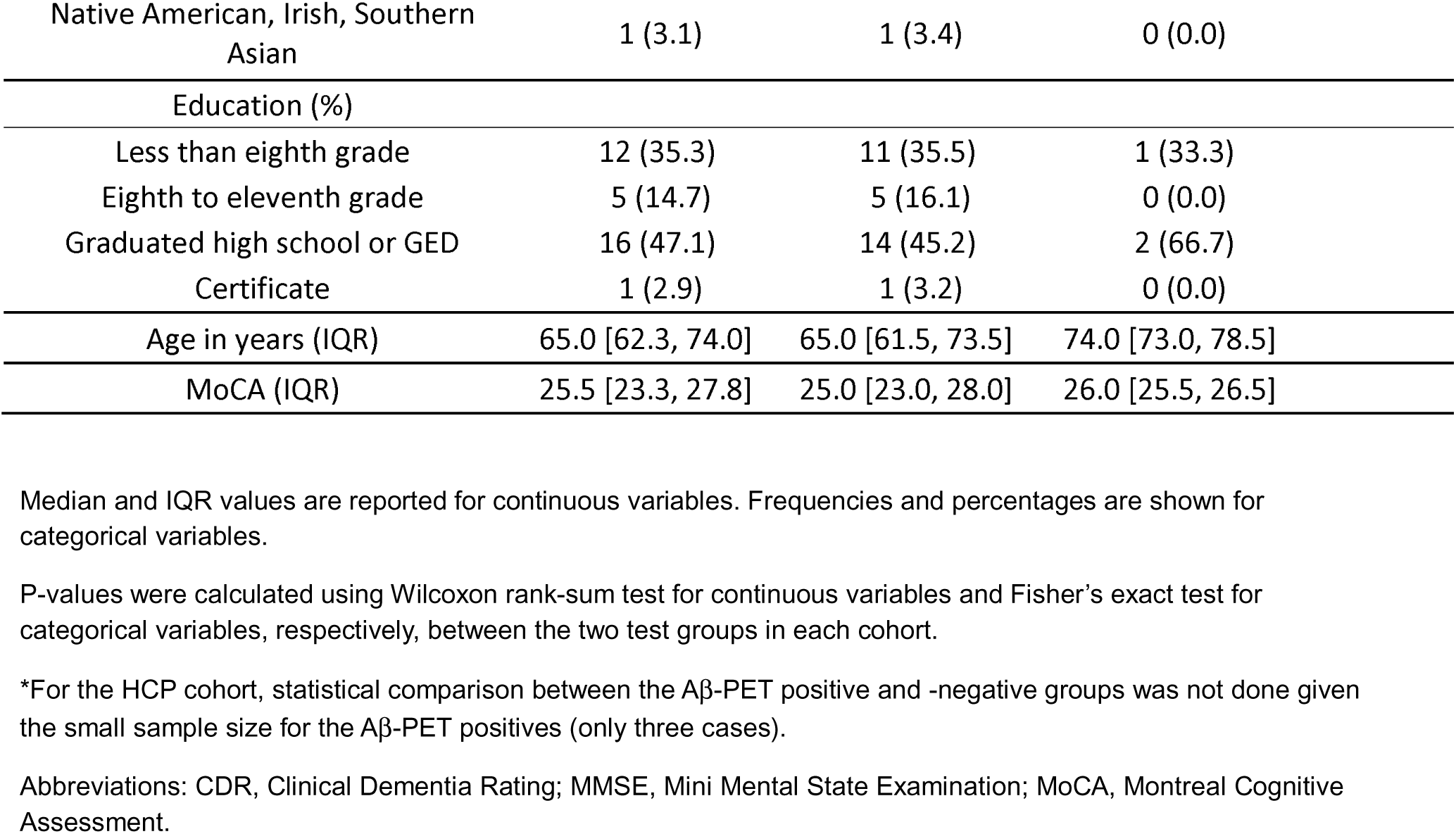
Demographic and clinical characteristics of the Pitt-ADRC and Human Connectome Project cohorts.

| Pitt-ADRC Cohort | Overall | Normal control | Probable AD | p-value |
| --- | --- | --- | --- | --- |
| Overall | 50 | 34 | 16 |  |
| Sex (%) |  |  |  |  |
| Male | 29 (58.0) | 20 (58.8) | 9 (56.2) | 1.000 |
| Female | 21 (42.0) | 14 (41.2) | 7 (43.8) |  |
| CDR (%) |  |  |  |  |
| 0 | 16 (32.0) | 16 (47.1) | 0 (0.0) | <0.001 |
| 0.5 | 24 (48.0) | 16 (47.1) | 8 (50.0) |  |
| 1 | 5 (10.0) | 0 (0.0) | 5 (31.2) |  |
| 2 | 5 (10.0) | 2 (5.9) | 3 (18.8) |  |
| Age in years (IQR) | 76.0 [70.3, 82.0] | 77.0 [70.3, 82.0] | 75.5 [70.8, 78.3] | 0.811 |
| MMSE (IQR) | 27.0 [23.0, 29.0] | 28.0 [27.0, 29.0] | 21.5 [18.5, 23.3] | <0.001 |
| MoCA (IQR) | 23.0 [16.5, 27.0] | 27.0 [23.0, 28.0] | 16.0 [14.0, 19.3] | <0.001 |

| Pitt-ADRC sub-cohort for NULISA | Overall | Normal control | Probable AD | p-value |
| --- | --- | --- | --- | --- |
| Overall | 43 | 29 | 14 |  |
| Sex (%) |  |  |  |  |
| Male | 25 (58.1) | 17 (58.6) | 8 (57.1) | 1.000 |
| Female | 18 (41.9) | 12 (41.4) | 6 (42.9) |  |
| CDR (%) |  |  |  |  |
| 0 | 15 (34.9) | 15 (51.7) | 0 (0.0) | 0.001 |
| 0.5 | 19 (44.2) | 12 (41.4) | 7 (50.0) |  |
| 1 | 4 (9.3) | 0 (0.0) | 4 (28.6) |  |
| 2 | 5 (11.6) | 2 (6.9) | 3 (21.4) |  |
| Age in years (IQR) | 76.0 [70.0, 81.5] | 77.0 [70.0, 82.0] | 75.5 [70.3, 76.8] | 0.755 |
| MMSE (IQR) | 27.0 [23.0, 29.0] | 29.0 [27.0, 29.0] | 22.0 [19.3, 23.8] | <0.001 |
| MoCA (IQR) | 25.0 [17.0, 28.0] | 27.0 [25.0, 29.0] | 15.5 [14.0, 19.8] | <0.001 |

| HCP Cohort | Overall | Aβ PET-Negative | Aβ PET-Positive | p-value * |
| --- | --- | --- | --- | --- |
| Overall | 34 | 31 | 3 |  |
| Sex (%) |  |  |  |  |
| Male | 10 (29.4) | 9 (29.0) | 1 (33.3) |  |
| Female | 24 (70.6) | 22 (71.0) | 2 (66.7) |  |
| Race (%) |  |  |  |  |
| White | 22 (68.8) | 19 (65.5) | 3 (100.0) |  |
| Black/African American | 9 (28.1) | 9 (31.0) | 0 (0.0) |  |

|  |  |  |  |
| --- | --- | --- | --- |
| Native American, Irish, Southern Asian | 1 (3.1) | 1 (3.4) | 0 (0.0) |
| Education (%) |  |  |  |
| Less than eighth grade | 12 (35.3) | 11 (35.5) | 1 (33.3) |
| Eighth to eleventh grade | 5 (14.7) | 5 (16.1) | 0 (0.0) |
| Graduated high school or GED | 16 (47.1) | 14 (45.2) | 2 (66.7) |
| Certificate | 1 (2.9) | 1 (3.2) | 0 (0.0) |
| Age in years (IQR) | 65.0 [62.3, 74.0] | 65.0 [61.5, 73.5] | 74.0 [73.0, 78.5] |
| MoCA (IQR) | 25.5 [23.3, 27.8] | 25.0 [23.0, 28.0] | 26.0 [25.5, 26.5] |

Median and IQR values are reported for continuous variables. Frequencies and percentages are shown for categorical variables.
P-values were calculated using Wilcoxon rank-sum test for continuous variables and Fisher's exact test for categorical variables, respectively, between the two test groups in each cohort.
\*For the HCP cohort, statistical comparison between the Aβ-PET positive and -negative groups was not done given the small sample size for the Aβ-PET positives (only three cases).
Abbreviations: CDR, Clinical Dementia Rating; MMSE, Mini Mental State Examination; MoCA, Montreal Cognitive Assessment.

Due to biospecimen limitations, we included a slightly smaller Pitt-ADRC cohort for the NULISA assay (n=43, instead of 50). The median age was 76 (70.0-81.5) years, with 18 (41.9%) females. Fourteen (32.6%) were diagnosed with probable AD and twenty-nine (69.4%) as normal controls. Cognitively, the median MMSE and MoCA scores were 27.0 (23.0- 29.0) and 25.0 (17.0- 28.0), respectively. Regarding CDR scores, 15 participants (34.9%) were CDR=0, 19 (44.2%) had CDR=0.5, 4 (9.3%) were CDR=1, and 5 (11.6%) had CDR=2. Comparing these metrics between the probable AD and normal control groups indicated significant differences in MoCA, MMSE and CDR scores. (Table 1)

In the HCP cohort (n=34), the median age was 65 (62.3-74.0) years, with 24 (70.6%) females. Three (8.8%) were Aβ-PET positive and thirty-one (91.2%) were Aβ-PET negative. The median MoCA score was 25.5 (23.3- 27.8). Regarding self-identified race, 22 participants (68.8%) were non-Hispanic White, 9 (29.1%) were African Americans, and one (3.1%) was native American, Irish or Southern Asian. For education, 12 (35.3%) had less than eighth grade education, 5 (14.7%) had eighth to eleventh grade education, 16 (47.1%) had graduated high school or general educational development (GED) education, and 1 (2.9%) had certificate education. (Table. 1)

### 3.2 Precision and detectability of p-tau217 in plasma versus serum

The precision of the assays, defined as the consistency between duplicate measures, was evaluated. The ALZpath, Lumipulse, and Pittsburgh assays demonstrated slightly better precision in serum (9.4%, 7.6%, and 9.2%, respectively) compared with plasma (10.7%, 10.1%, and 11.1%, respectively). Precision for p-tau217 in the NULISA assay was not measured, as all samples were analyzed in a single run.

For detectability, the ALZpath assay had two serum samples below the low limit of quantification (LLOQ) of 0.00978 pg/mL. In contrast, all samples analyzed with the Lumipulse assay had concentrations above its LLOQ of 0.037 pg/mL. [25] For NULISA, one plasma sample and six serum samples were below the target LLOQ (2^NPQ=1625.52).

Together, p-tau217 exhibited slightly better precision in serum versus plasma, while detectability of ALZpath and NULISA appeared more favorable in plasma than serum.

### 3.3 Concentration difference between matched plasma and serum

P-tau217 concentrations (pg/mL) were compared between plasma and serum across all assays. In the Pitt-ADRC cohort, both the ALZpath (plasma = 0.47 [0.31–0.91] pg/mL vs. serum = 0.35 [0.19–0.63] pg/mL) and Pittsburgh (plasma = 25.0 [17.95–57.06] pg/mL vs. serum = 18.38 [10.61–33.90] pg/mL) assays showed significantly higher (p < 0.0001) p-tau217 levels in plasma compared with serum. Conversely, the Lumipulse assay demonstrated significantly higher (p < 0.0001) levels in serum (0.27 [0.18–0.70] pg/mL) than in plasma (0.26 [0.15–0.59] pg/mL) (Figure 1A, B). A similar trend was observed in the HCP cohort, where the ALZpath assay showed significantly higher (p < 0.0001) p-tau217 levels in plasma (0.21 [0.15–0.29] pg/mL) than in serum (0.08 [0.05–0.11] pg/mL), and the Pittsburgh assay gave 8.79 [7.33–15.26] pg/mL in plasma compared with 4.29 [2.17–10.11] pg/mL in serum. However, the Lumipulse assay showed no significant difference between plasma (0.11 [0.09–0.17] pg/mL) and serum (0.12 [0.09–0.15] pg/mL) in the HCP cohort. (Figure 2A, B)

**Figure 1:**
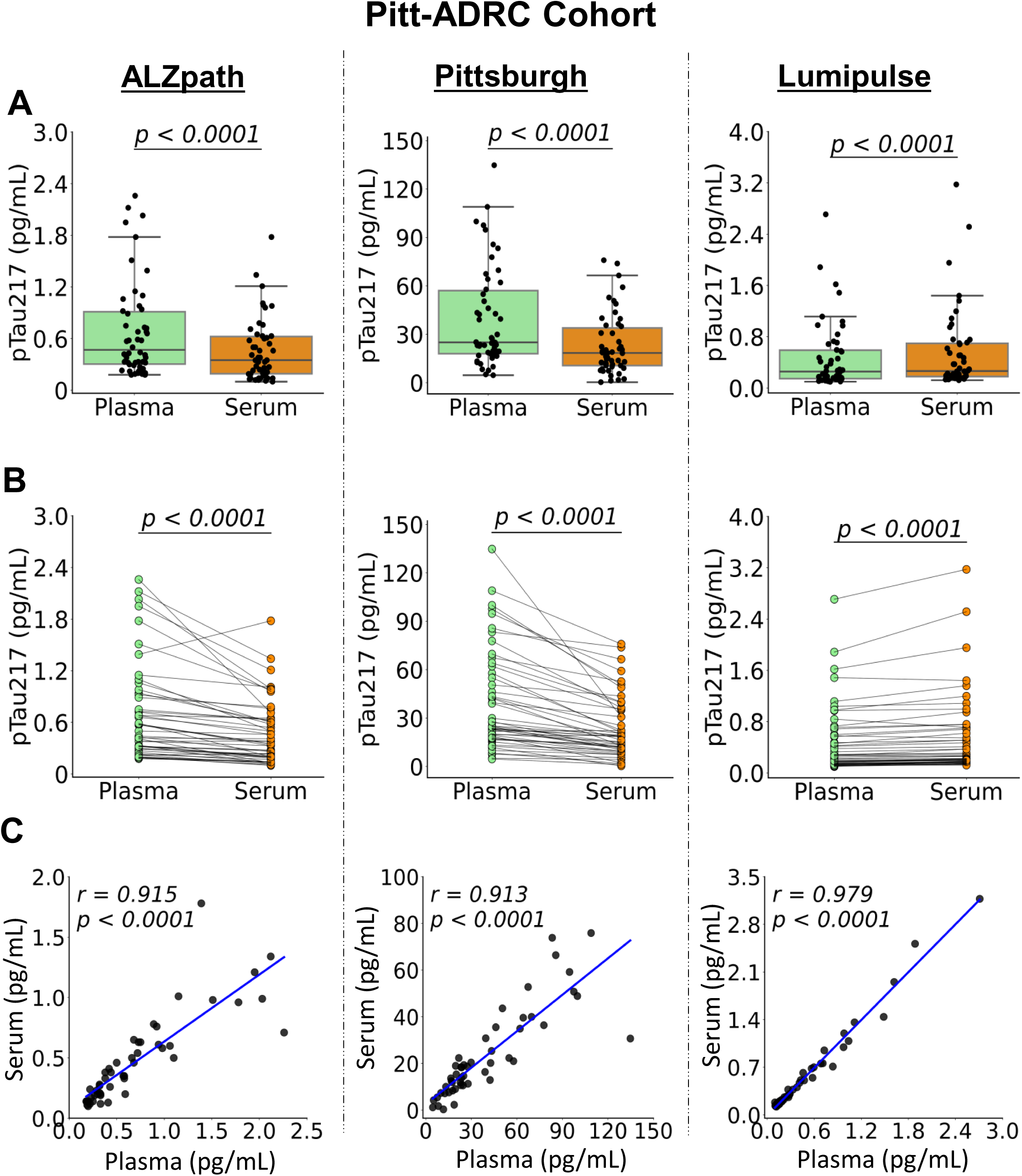
Comparison of p-tau217 levels between matched plasma and serum samples in the Pitt-ADRC cohort for three assays: ALZpath, Pittsburgh, and Lumipulse. **A.** Boxplots distribution of p-tau217 concentrations. The center line of each box represents the median, the edges are the 25th and 75th percentile, and the whiskers extend to the furthest non-outlier data value (1.5x IQR away from the 25th and 75th percentiles). **B.** Scatter plot distributions with lines connecting matched plasma and serum samples from the same individual. P values indicate the significance of the difference between plasma and serum according to the Wilcoxon signed rank test in concentrations. **C.** Correlation plots between plasma and serum concentrations. Correlation coefficients and associated p values were based on Spearman correlation. Lines represent the linear regression line. N= 50 for the Pitt-ADRC cohort across all assays.

**Figure 2:**
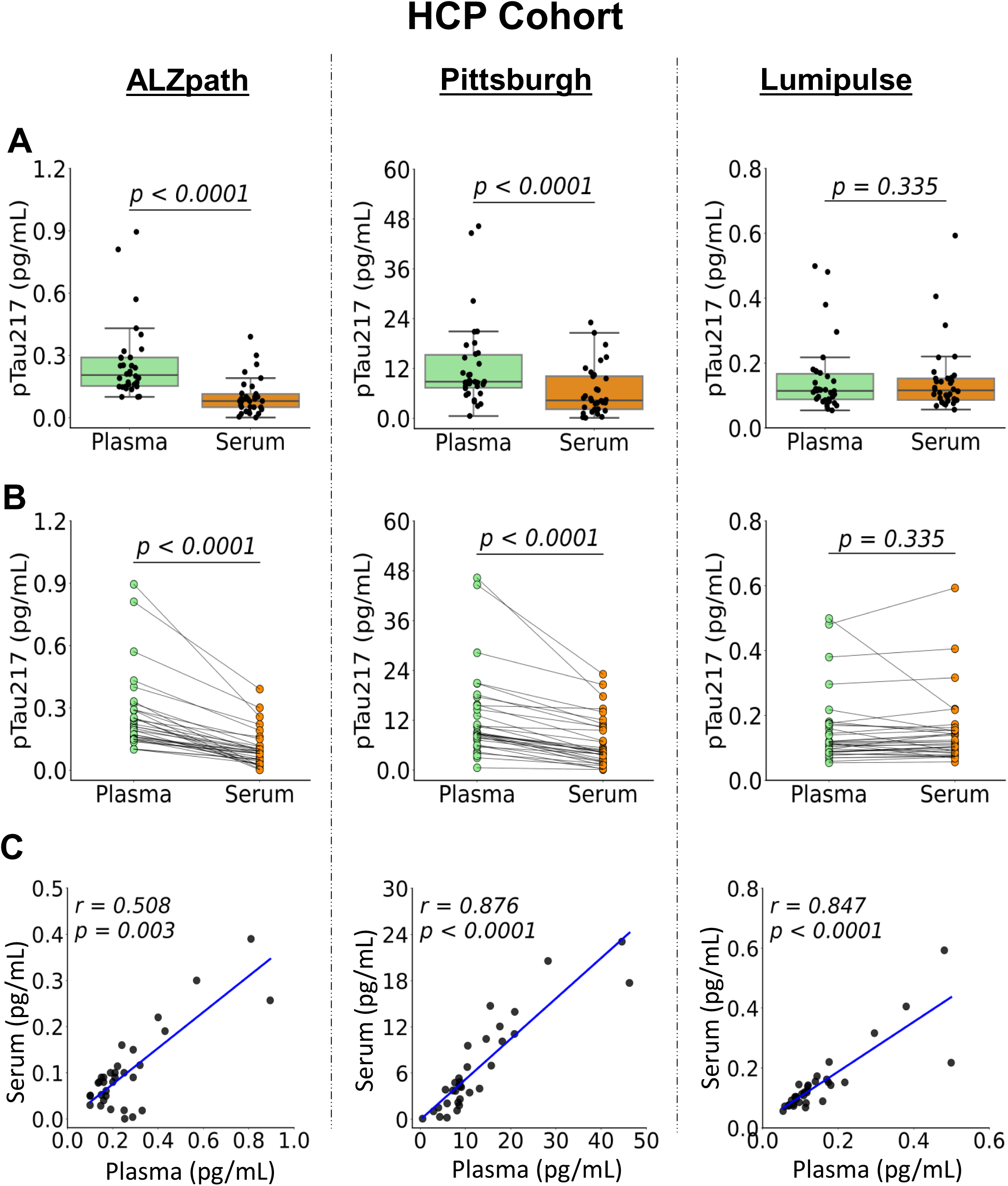
Comparison of p-tau217 biomarker concentrations between matched plasma and serum samples in the HCP cohort for three assays: ALZpath, Pittsburgh, and Lumipulse. **A.** Boxplots depicting the distribution of p-tau217 concentrations in plasma and serum for the three assays. The center line of each box represents the median, the edges are the 25th and 75th percentile, and the whiskers extend to the furthest non-outlier data value (1.5x IQR away from the 25th and 75th percentiles). **B.** Scatter plots with lines connecting plasma and serum samples from the same individual. P values were according to the Wilcoxon signed rank test. **C.** Correlation plots between plasma and serum concentrations for each assay Correlation coefficients and associated p values were based on Spearman correlation. Lines represent the linear regression lines. N=34 for Lumipulse and n=33 for Pittsburgh and ALZPath.

Plasma and serum p-tau217 in the Pitt-ADRC cohort showed strongly positive correlations. The strongest correlation was observed for the Lumipulse assay with r=0.979. (Figure 1C) while ALZpath and Pittsburgh had r=0.915 and r=0.913, respectively. In the HCP cohort, Lumipulse and Pittsburgh recorded high correlations (r=0.847 and r=0.876, respectively), while ALZpath displayed a moderate correlation (r=0.508). (Figure 2C)

For NULISA, there were significantly higher (p<0.0001) p-tau217 concentrations in plasma 3191.46 [2164.98-5367.37] 2^NPQ) than in serum (2486.67 [1728.23-3492.39] 2^NPQ). (Figure 3A, B) Correlation analysis revealed a strong positive relationship between plasma and serum (r=0.902). (Figure 3C).

**Figure 3:**
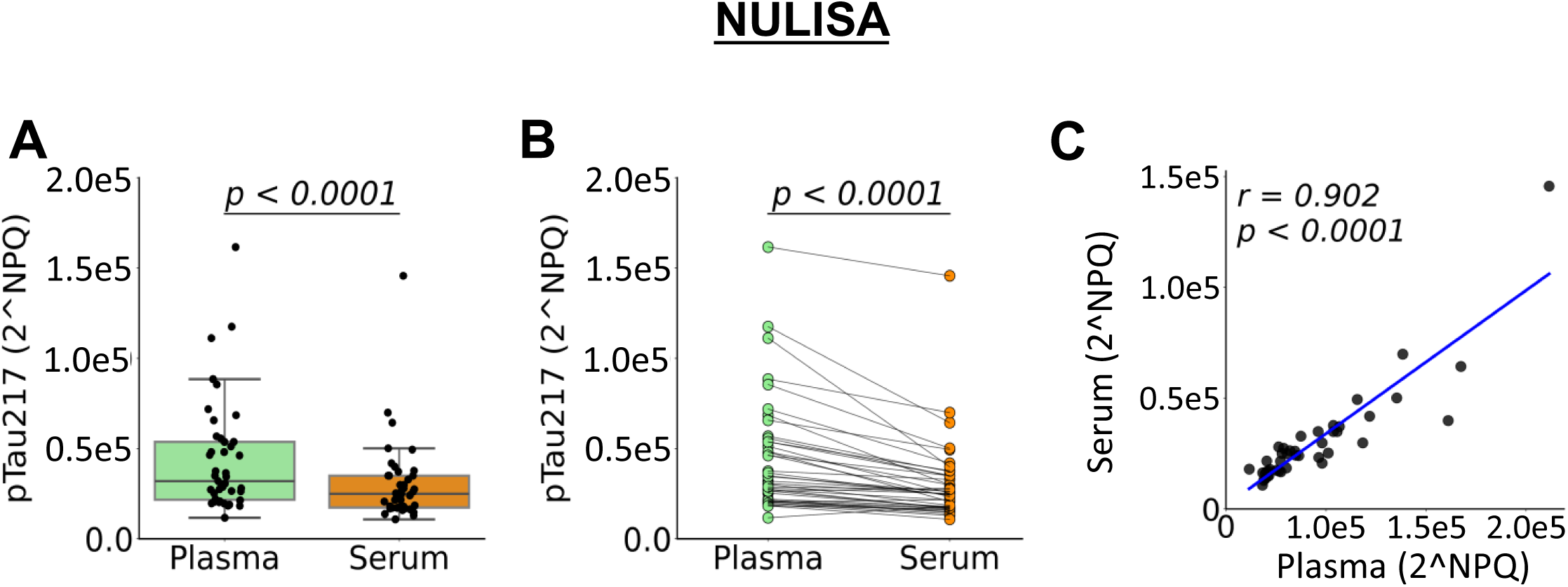
Comparison of p-tau217 concentration values between matched plasma and serum samples measured with the NULISA assay. **A.** Boxplots depicting the distribution of inversed log2 NULISA Protein Quantification (2^NPQ) values in plasma vs. serum. The center line of each box represents the median, the edges are the 25th and 75th percentile, and the whiskers extend to the furthest non-outlier data value (1.5x IQR away from the 25th and 75th percentiles). **B.** Individual NPQ values for matched plasma and serum are connected by lines to illustrate paired comparisons for identical participants. **C.** Correlation plots between plasma and serum NPQ values for each assay analyzed using Spearman correlation. P-values indicating statistical significance (p < 0.0001). N=43 for the NULISA using Pitt-ADRC reduced cohort.

These results show that p-tau217 levels were higher in plasma than matched serum for the Pittsburgh, ALZpath and NULISA assays. However, Lumipulse recorded similar/equivalent p-tau217 levels when measured in either matrix, showing matrix-based differences for the various assays.

### 3.4 Clinical performance of p-tau217 using different sample types across assays

Plasma and serum p-tau217 were evaluated in distinguishing clinically diagnosed AD status from normal controls. The ALZpath, Pittsburgh, NULISA and Lumipulse assays all had higher p-tau217 concentrations for probable AD individuals compared with normal controls across all assays (p<0.0001), whether plasma or serum was used. (Figure 4A)

**Figure 4:**
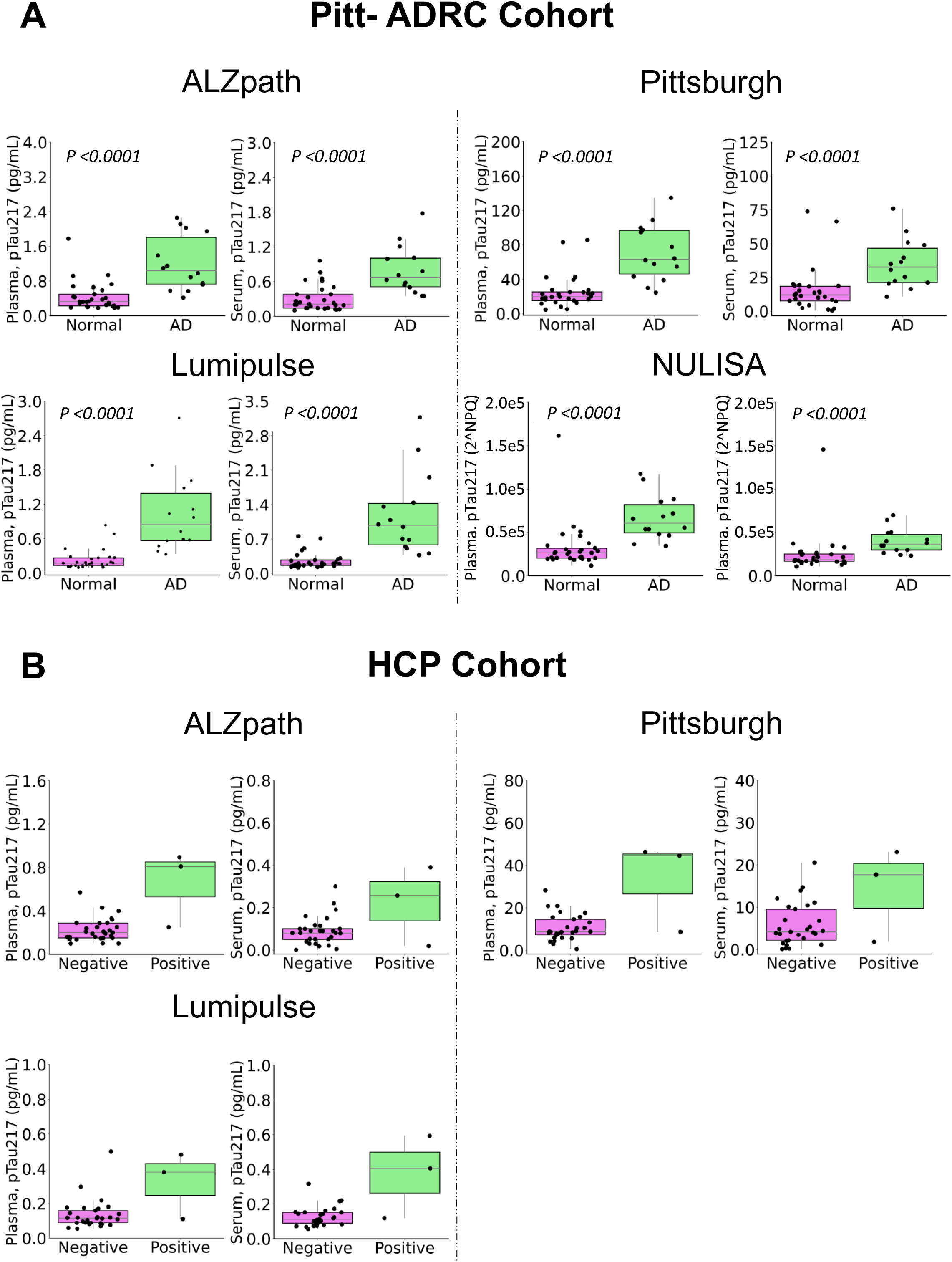
The boxplots illustrate the clinical performance of p-tau217 in matched plasma and serum samples for the Pitt-ADRC cohort (according to clinical diagnosis; A) and the HCP cohort (by **A**β PET status; B). The center line of each box represents the median, the edges are the 25th and 75th percentile, and the whiskers extend to the furthest non-outlier data value (1.5x IQR away from the 25th and 75th percentiles). The p-values were calculated using Wilcoxon rank sum test for the Pitt-ADRC cohort. Due to having only three cases in the Aβ PET positive sample size, no statistical analysis was performed for the HCP cohort. The normal control (Normal) and Probable AD (AD) were assessed through clinical assessment. The Negative and Positive were assessed based on Aβ PET imaging result.

The Area Under the Curve (AUC) results for p-tau217 to separate probable AD from normal controls demonstrated high performance across all methods. For Lumipulse, both plasma (AUC=0.963) and serum (AUC=0.961) p-tau217 showed similarly high accuracies. Furthermore, the ALZpath assays showed high reliability in both plasma and serum with AUCs of 0.916 and 0.899, respectively. The Pittsburgh assay achieved AUCs of 0.912 in plasma and 0.844 in serum, suggesting better performance in plasma. This was similar to NULISA with an AUC of 0.918 for plasma and 0.879 for serum. The Delong test between the corresponding plasma and serum samples indicated no difference in performance for all assays except for the Pittsburgh method (p=0.014, Supplement table 1). (Figure 5)

**Figure 5:**
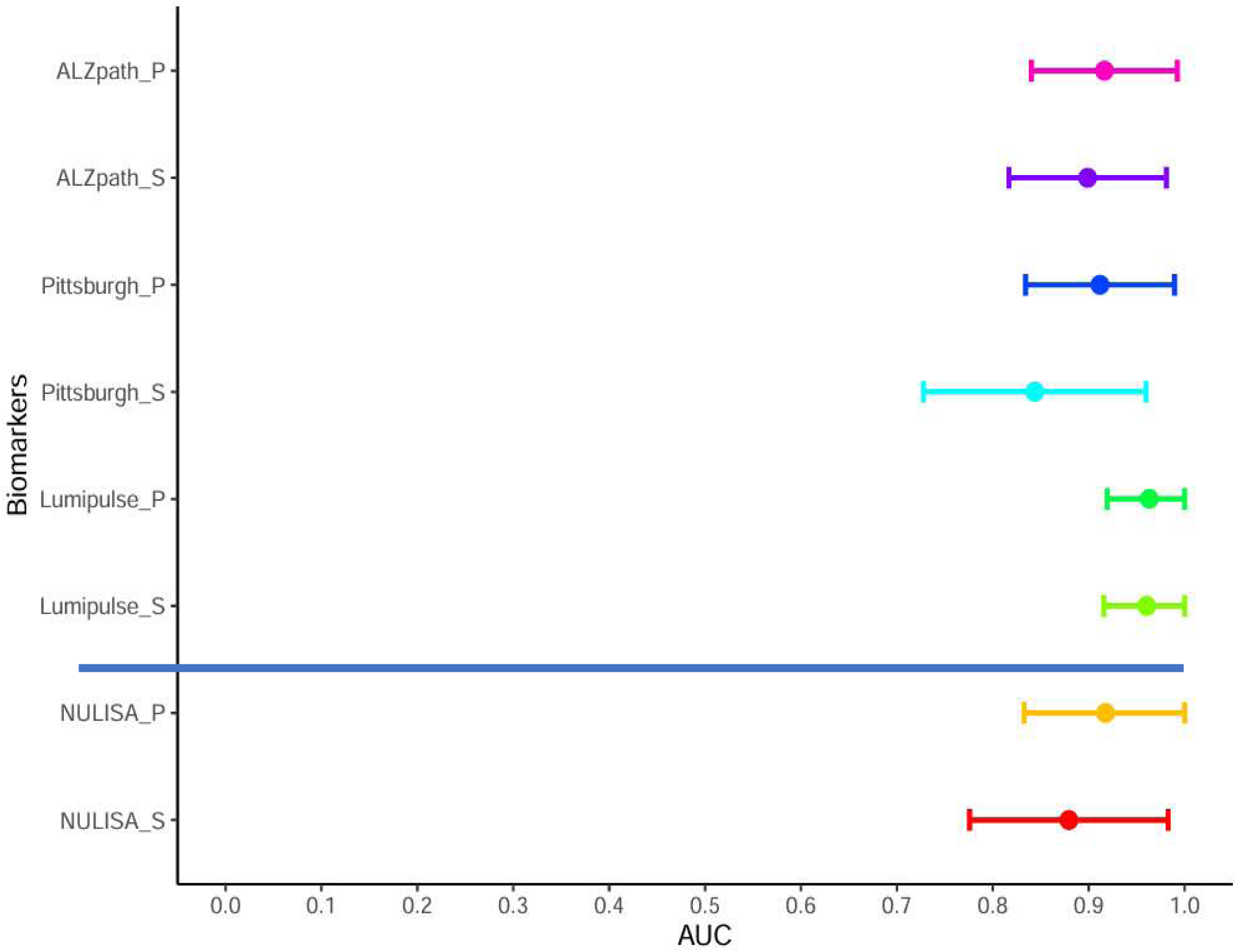
Performance of serum vs. plasma p-tau217 to differentiate individuals with probable AD from normal controls in the Pitt-ADRC cohort. The figure illustrates the AUC and 95% confidence interval of the assays. “P” represents plasma and “S” represents serum. Calculations of the AUC were based solely on the Pitt-ADRC cohort (n=50). The NULISA assays were conducted using the same cohort but with a reduced number of samples (n=43).

In the HCP cohort, p-tau217 levels in both plasma and serum were higher in the Aβ-PET positive versus Aβ-PET negative groups. However, due to the limited sample size in the Aβ-PET positive group (only 3 cases), statistical testing or AUC comparisons were not performed (Figure 4B).

The results indicated equivalent clinical performance of p-tau217 assays in plasma and serum for Lumipulse, ALZpath, and NULISA. However, the Pittsburgh assay demonstrated significantly better performance in plasma compared to serum.

### 3.5 Correlation between different assay platforms

In Pitt-ADRC, all assay comparisons showed significant correlations. Plasma measurements indicated strong correlations with the r-value of 0.848 for Pittsburgh vs. ALZpath, 0.915 for Lumipulse vs. ALZpath, and 0.868 for Lumipulse vs. Pittsburgh. Serum measurements displayed relatively moderate correlations, with correlation coefficients of 0.752 for Pittsburgh vs. ALZpath, 0.892 for Lumipulse vs. ALZpath, and 0.762 for Lumipulse vs. Pittsburgh. Complete correlation information is shown in Supplementary Figure 1A.

In the reduced Pitt-ADRC cohort, NULISA also showed strong correlations with the other assays. In plasma, the correlation coefficients were 0.785 against ALZpath, 0.793 against Pittsburgh, and 0.888 against Lumipulse. In serum, the correlation coefficients were 0.799 against ALZpath, 0.756 against Pittsburgh, and 0.857 against Lumipulse. (Supplementary Figure 1B)

In HCP, significant correlations were also observed across all assays. Plasma measurements demonstrated moderate to strong correlations, with correlation coefficients of 0.648 for Lumipulse vs. ALZpath, 0.828 for Pittsburgh vs. ALZpath, and 0.632 for Lumipulse vs. Pittsburgh. Serum measurements showed lower correlations, following a similar trend, with correlation coefficients of 0.357 for Lumipulse vs. ALZpath, 0.505 for Lumipulse vs. Pittsburgh, and 0.435 for Pittsburgh vs. ALZpath. (Supplementary Figure 1C)

Together, p-tau217 assays from different sources show moderate to strong correlations when measured in either plasma or serum.

## Discussion

Plasma has been the primary matrix for p-tau217 measurements and has already been head-to-head compared across multiple assays. [26] Due to its broader availability in clinical practice, serum represents a critical alternative matrix for p-tau217 measurement. [17] Establishing the reliability of serum is particularly important as p-tau217 continues to gain prominence in the AD field. In this study, a pioneering comparison of p-tau217 levels in plasma and serum was conducted across four distinct assays. Our findings revealed notable differences in p-tau217 levels between matched plasma and serum samples across most but not all assays. Specifically, p-tau217 levels were lower in serum vs. plasma for the ALZpath, Pittsburgh, and NULISA assays, while Lumipulse p-tau217 showed high levels in serum that reached significance in one cohort. These results may be attributed to potential differences in assay binding characteristics, and differences in assay design and/or protein recovery efficiency between serum and plasma. [27–29]

All p-tau217 assays examined exhibited high clinical performance in distinguishing between probable AD and normal controls when serum in lieu of plasma was used. The AUC values in the Pitt-ADRC cohort was 0.84-0.96, and statistical analyses revealed no significant differences in AUC between the plasma and corresponding serum samples for the Lumipulse, ALZpath, and NULISA p-tau217 assays, establishing a potential to use serum for p-tau217 measurements using these assays. These results show serum p-tau217 as a feasible option in both research and clinical settings.

While the clinical performance of most p-tau217 assays was comparable between serum and plasma, allowing either matrix to be used in clinical and research settings, it is crucial to avoid intermixing plasma and serum in studies, such as in longitudinal analyses, due to variability in absolute concentrations between the two matrices. Thus, plasma and serum must be used independently for any given study.

We did not observe significant differences in precision between the Lumipulse, ALZpath, and Pittsburgh assays. However, both ALZpath and NULISA recorded some serum samples that fell below the LLOQ. By extension, this suggests that more serum samples are likely to return outside the measurable zone when using these assays. Thus, serum might not be preferrable for evaluating predominantly cognitively normal populations where p-tau217 levels tend to be much lower than in cognitively impaired individuals. [9, 10, 15, 30]

While our results show that all the evaluated platforms can be used for serum p-tau217 measurement, the method selection must be carefully considered. The Lumipulse assay showed similar or higher levels of p-tau217 in serum relative to plasma, thereby reducing concerns of sample type-dependent detectability issues. Moreover, the Lumipulse platform, which is FDA-approved for *in vitro* diagnostic use for CSF Aβ and tau biomarker assays and is already being considered by the FDA for plasma p-tau217 approval, might have potential applicability in broader biomarker contexts. Nevertheless, the Lumipulse assay requires a higher dead volume, and a higher sample volume compared with the other assays, limiting its utility in scenarios with restricted sample volume availability. In such cases, the ALZpath, Pittsburgh, or NULISA assays may be more suitable for serum p-tau217 measurement. On the other hand, the NULISA platform offers the capability to measure over 100 biomarkers simultaneously, presenting a valuable tool for comprehensive biomarker studies. However, its higher cost and inability to provide absolute quantification at its current stage may limit its suitability for clinical studies at the current time. The ALZpath and Pittsburgh assays, both utilizing the HD-X Simoa instrument, offer versatility for defined-assay workflows, making them suitable for targeted serum p-tau217 analyses.

Our findings on p-tau217 share similarities with those from a previous study on plasma p-tau181 and p-tau231, with the two biomarkers having higher absolute levels in plasma than serum but equivalent accuracies to differentiate Aβ-positive AD dementia from Aβ-negative controls. [31]

### Strengths and limitations

Our study has several significant strengths. First, we included two separate cohorts – one from a memory clinic setting and the other from a racially diverse community-based research study, enhancing the generalizability and practical relevance of our findings due to their diverse representation. Moreover, while our analysis of the Pitt-ADRC cohort focused on cognitive status, our interest in the HCP cohort was aimed at brain Aβ pathology according to PiB PET imaging. Each cohort was thoroughly characterized by clinical evidence of disease through cognitive evaluations including the MMSE, MoCA, and CDR, as well as clinical assessment or brain Aβ PET imaging. Additionally, we evaluated p-tau217 using four different assays, providing a comprehensive analysis of the diagnostic utility of serum p-tau217 across various platforms. This multi-faceted approach ensures a robust examination of p-tau217 as a potential biomarker for AD.

Limitations of this study include the following. Firstly, the sample size across cohorts was relatively small, particularly for the NULISA assay, where only one cohort with a reduced sample size was processed. This limitation may affect the generalizability of our results. Future evaluations utilizing a larger scale cohort of participants across the AD continuum will be needed to confirm our findings. Furthermore, the HCP cohort contained limited numbers of Aβ PET-positive patients. Further validation using a larger cohort with a substantial number of Aβ PET-positive individuals will be necessary.

## Conclusion

We evaluated serum p-tau217 using four assays and compared the results with plasma p-tau217 levels in matched samples. Significant correlations between plasma and serum p-tau217 were observed within all assays. Both plasma and serum p-tau217 showed strong AD diagnostic performance across the assays, however the absolute levels showed matrix-dependent variations. This study offers comprehensive insights into serum p-tau217 and its measurement, supporting its potential integration into clinical settings for early detection and management of AD. Additionally, it provides insights into assay-to-assay differences in performance, aiding in the selection of appropriate assays for clinical and research applications.

## Non-standard abbreviations

AD: Alzheimer disease
p-tau: phosphorylated tau
EDTA: ethylenediaminetetraacetic acid
AUC: area under the curve
PET: positron emission tomography
CSF: cerebrospinal fluid
p-tau217: phosphorylated tau217
Aβ: amyloid-beta
NULISA: nucleic acid linked immuno-sandwich assay
Pitt-ADRC: University of Pittsburgh Alzheimer’s Disease Research Center
MoCA: Montreal Cognitive Assessment
MMSE: Mini-Mental State Examination
CDR: Clinical Dementia Rating
HCP: Human Connectome Project
PiB: Pittsburgh Compound B
MRI: magnetic resonance imaging
Simoa: single molecule array
IQR: interquartile range
ROC: receiver operating characteristic
NPQ: NULISA Protein Quantification
2^NPQ: log2 transformation
GED: general educational development
LLOQ: lower limit of quantification.

## Author contributions

The corresponding author takes full responsibility that all authors on this publication have met the following required criteria of eligibility for authorship: (a) significant contributions to the conception and design, acquisition of data, or analysis and interpretation of data; (b) drafting or revising the article for intellectual content; (c) final approval of the published article; and (d) agreement to be accountable for all aspects of the article thus ensuring that questions related to the accuracy or integrity of any part of the article are appropriately investigated and resolved. Nobody who qualifies for authorship has been omitted from the list.

## Research funding

The Pittsburgh ADRC and the HCP study were funded by NIH/NIA grants P30AG066468 and R01AG072641 respectively. TKK and the Karikari Laboratory were supported by the NIH/NIA grants R01AG083874, U24AG082930 and P01AG025204, and a professorial endowment from the Department of Psychiatry, University of Pittsburgh.

## Data Availability

All data produced in the present study are available upon reasonable request to the authors

## Acknowledgements

We thank the Pittsburgh ADRC and HCP study participants and their families and caregivers.

