## Supplementary material for "Equivalence of plasma and serum for clinical measurement of p-tau217: comparative analyses of four blood-based assays": Supplymentary

Supplement Figure 1

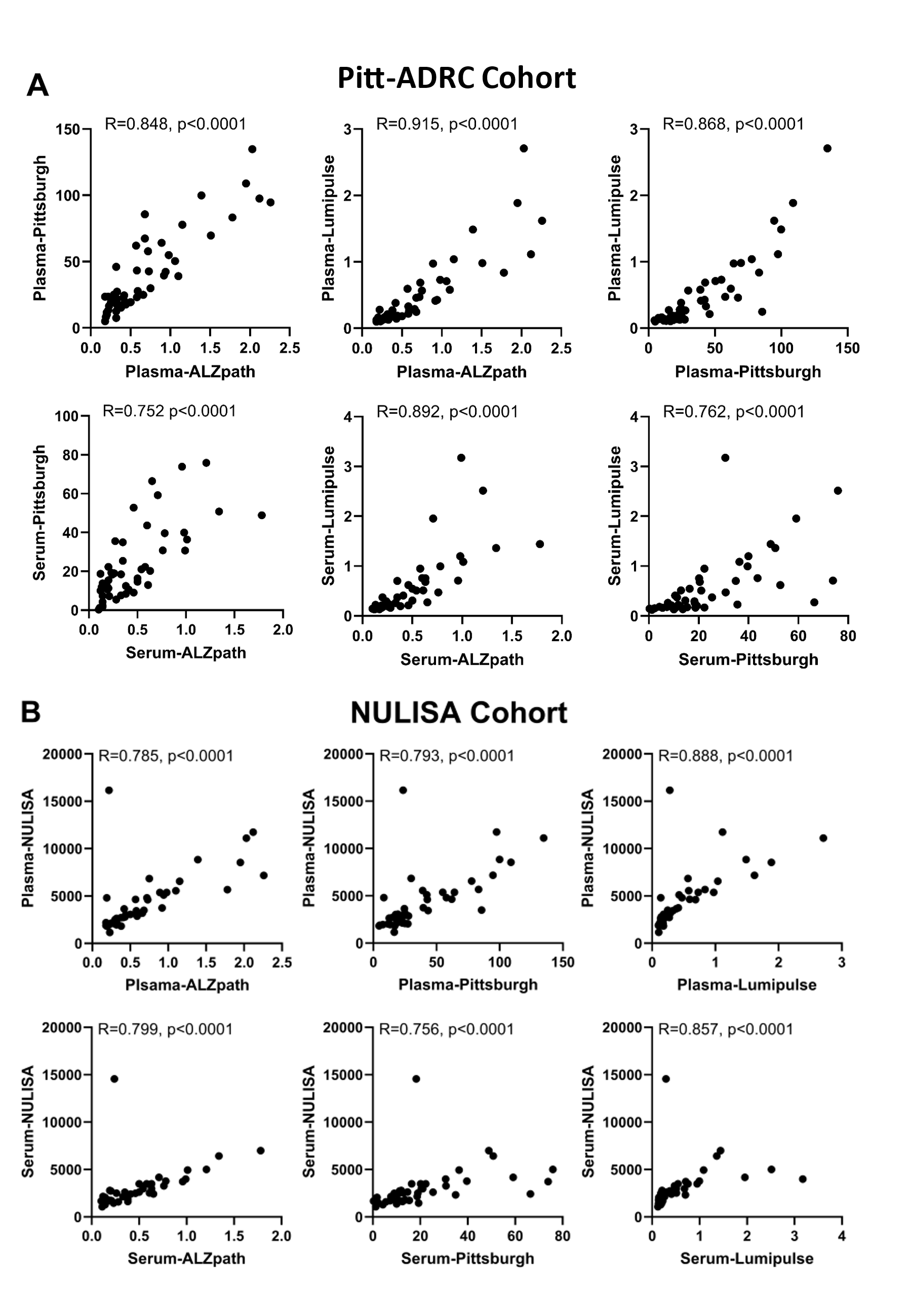

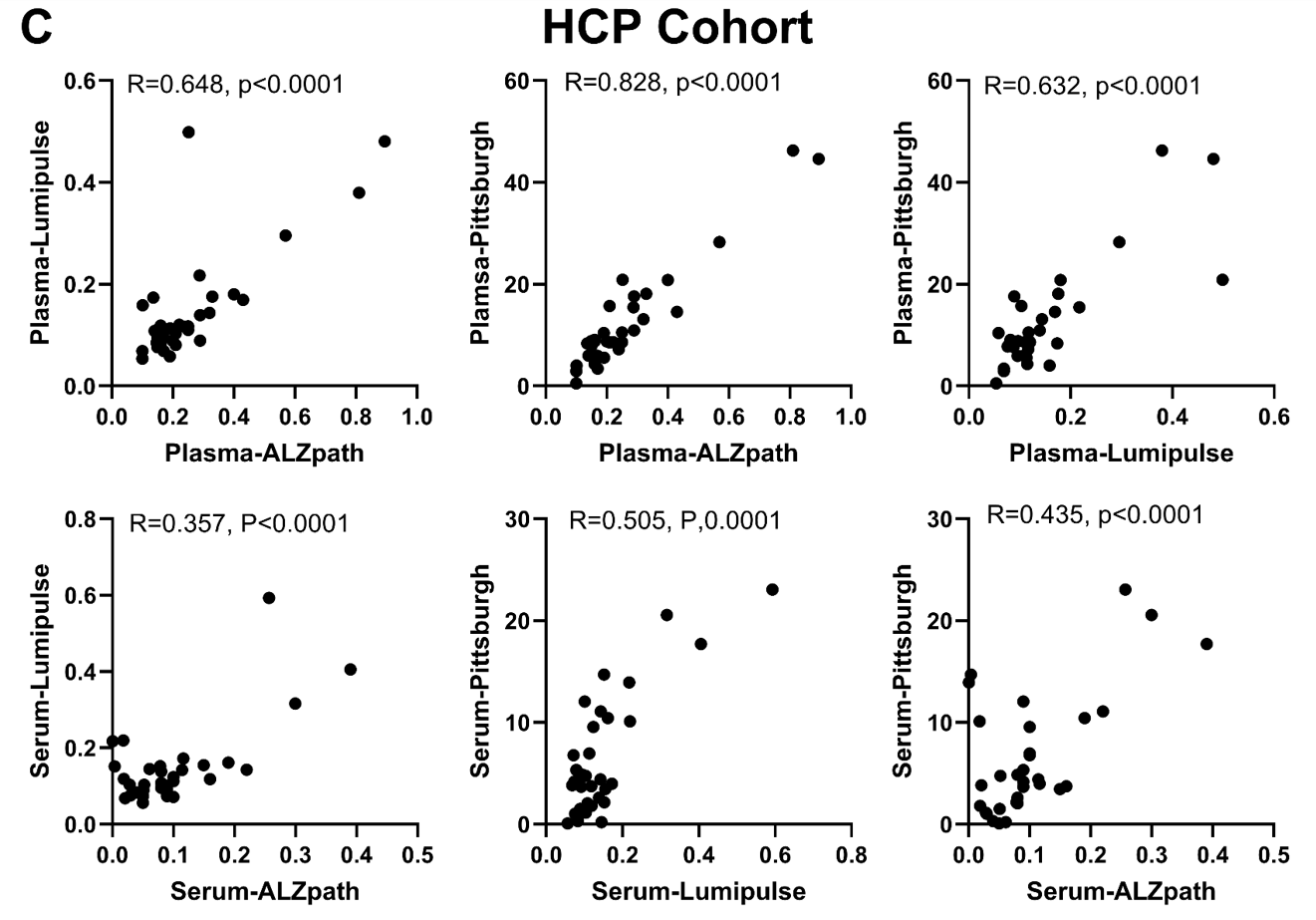
 **Supplement Figure 1: Between-assay correlation of plasma or serum p-tau217 in the Pitt-ADRC and HCP cohorts.** The figure illustrates the correlation between different assays for Pitt-ADRC cohort (A), NULISA reduced cohort (B) and HCP cohort (C). The analysis utilizes the Spearman test to evaluate the relationships between them.

Supplement Table 1. Results of the Delong test comparing the AUCs of corresponding plasma and serum assays

| DeLong test result | | |
| --- | --- | --- |
| Plasma | Serum | p-value |
| ALZpath_P | ALZpath_S | 0.452 |
| Pittsburgh_P | Pittsburgh_S | 0.014 |
| Lumipulse_P | Lumipulse_S | 0.716 |
| NULISA_P | NULISA_S | 0.197 |
